# “Prevention is better than cure” - A Mixed Methods Study of Communities’ and Healthcare Workers’ Perspectives on New Adult and Adolescent TB Vaccines and their Implementation in Zambia

**DOI:** 10.1101/2025.07.15.25331585

**Authors:** Andrew D. Kerkhoff, Jake M. Pry, Mwiza Haambokoma, Noelle Le Tourneau, Sandra S. Simbeza, Bertha Shamoya, Nelly Zulu, Herbert Nyirenda, Evelyn Kundu-Ng’andu, Kombatende Sikombe, Elvin H. Geng, Rupali J. Limaye, Anjali Sharma

## Abstract

**Background:** New tuberculosis (TB) vaccines for adults and adolescents are crucial for achieving global control targets, and several candidates may be ready for introduction within five years. We conducted a convergent mixed-methods study in Lusaka, Zambia, among community members and healthcare workers (HCWs) to understand preliminary TB vaccine acceptance and potential factors influencing uptake to inform readiness activities critical for successful implementation.

**Methods:** Adult community members were enrolled from randomly selected households within four communities with historically low COVID-19 vaccine coverage, and HCWs from 10 public healthcare facilities representing different care levels. Structured surveys evaluated participants’ perspectives on new vaccines, and mixed-effects Poisson regression was used to estimate the marginal probability of participants’ intention to get a new TB vaccine. Qualitative in-depth interviews (IDIs; n=25) and focus group discussions (FGDs; n=9) were conducted and analyzed using a hybrid approach to explore acceptable vaccine attributes, preferences, and determinants of TB vaccine uptake, and recommendations for rollout.

**Results:** Overall, 499 participants completed surveys (395 community members, 104 HCWs), with 63 additional participants across IDIs and FGDs. 77% of community members and 83% of HCWs expressed intention to get a TB vaccine. Perceived TB risk and severity concerns were the only significant drivers of vaccine intention. Participants pragmatically accepted WHO minimum vaccine specifications (50% efficacy, two-dose schedule, transient mild side effects, at least 2 years duration), though HCWs preferred higher efficacy thresholds due to occupational risk. Implementation preferences emphasized early community engagement (at least 3-6 months pre-rollout), diverse delivery approaches including facility-based and community venues for adults, schools for adolescents, and door-to-door campaigns, while addressing COVID-19-related skepticism through transparent and clear communication using trusted sources.

**Conclusions:** This study reveals strong potential for successful TB vaccine rollout in Zambia through proactive implementation strategies, including early community engagement, transparent communication, and person-centered delivery approaches.

## Background

Models indicate that failure to introduce a new tuberculosis (TB) vaccine could result in an additional 26.5 million cases, 2.8 million deaths, and nearly $500 billion in economic losses globally by 2030.^1^ This stark projection underscores why a new TB vaccine for adults and adolescents is critical to combat the ongoing global epidemic. Each year, more than 10.6 million individuals develop TB, contributing to substantial suffering and poor quality of life.^2^ TB remains the leading cause of death by infectious disease (over 1.3 million deaths annually) and is the leading cause of death among people with HIV (PWH).^2^ While current strategies to END TB by 2030 aim to reduce new cases and deaths by 80% and 90%, respectively, progress has been largely stagnant, partly due to the absence of a vaccine that effectively protects adults and adolescents.^1^ The century-old Bacillus Calmette–Guérin (BCG) vaccine provides virtually no protection to these age groups who are key to halting TB transmission,^3^ leaving a significant gap in available prevention tools.

The TB vaccine landscape is rapidly evolving, with several promising candidates, including M72/AS01E and MTBVAC, offering concrete hope for breakthrough prevention strategies within the next five years.^4^ However, as the COVID-19 pandemic demonstrated, scientific breakthroughs alone are insufficient for public health impact. Even highly efficacious vaccines can have limited impact when uptake is hindered by mistrust, misinformation, low-risk perception, and delivery challenges. COVID-19 experiences revealed both successful approaches and critical pitfalls: early vaccination of healthcare workers (HCWs) helped demonstrate safety and build community confidence, mobile clinics and workplace programs improved access for hesitant populations, and consistent messaging from trusted sources countered misinformation.^5–10^ Conversely, frequent changes to dosing recommendations, inconsistent supply, and inadequate community engagement undermined trust and reduced uptake in many settings.

With one or more TB vaccine candidates potentially ready for introduction within the next five years,^4^ the time to prepare for implementation is now. Incorporating the lessons from the COVID-19 pandemic and other recent vaccine introductions^5–13^ to ensure the impact of a new TB vaccine is optimized will require proactive planning, community engagement, and tailored implementation strategies informed by stakeholder perspectives. The World Health Organization (WHO) has developed a global framework to guide countries in planning, coordination, and readiness for new TB vaccine rollout, emphasizing early and comprehensive engagement with stakeholders to understand perspectives and preferences for country-specific readiness planning.^14^

Community members and HCWs are two critical stakeholder groups whose perspectives are fundamental to understanding and facilitating broad and equitable TB vaccine uptake. Community members represent the broader target population, while HCWs serve both as high-risk beneficiaries due to occupational exposure and as trusted advocates and intermediaries for community vaccination efforts. We conducted this mixed-methods study in Lusaka, Zambia, to explore community members’ and HCWs’ perspectives on new TB vaccines, including intentions, delivery preferences, and anticipated barriers to uptake. These findings serve to guide planning considerations, including targeted sensitization and communication strategies, to support the successful rollout of a future TB vaccine.^14,15^

## Methods

We employed a convergent, parallel mixed methods design to comprehensively understand TB vaccine attitudes, intentions, beliefs, and preferences among adult community members and HCWs. This approach allowed us to integrate quantitative (survey) and qualitative data (in-depth interviews (IDIs) and focus group discussions (FGDs)), providing a more nuanced exploration of perspectives related to new adult TB vaccines.

### Study Population and Setting

We conducted the study in Lusaka, Zambia in four urban neighborhoods, purposefully selected for comparatively low COVID-19 vaccine coverage, and at 10 healthcare facilities representing different levels of care (one tertiary teaching hospital, five first-level hospitals, three Urban Health Centers, and one Rural Health Centre).

Recruitment for quantitative data collection took place between 13 November 2023 and 15 December 2023 and closed once the pre-specified sample size was reached: 400 community members (100 per community) and 100 HCWs (10 from each facility) representing a mix of professional (doctor, nurse, clinical officer), and lay (community HCW, volunteer) HCWs.^16^ Recruitment for qualitative data collection took place between 4 April 2024 and 6 June 2024 and closed once the pre-specified number of FGDs was reached (N=9; 3 community members, 3 professional HCWs, 3 lay HCWs) and information saturation was reached for IDIs (N=25; 12 community members, 7 professional HCWs, 6 lay HCWs).

Community members were eligible if they were 18 years or older, resided in Lusaka Province, and provided informed consent in English or the two most commonly spoken non-English languages in Zambia, Bemba or Nyanja. HCWs were additionally required to provide direct clinical or community-based health services.

Research teams for the survey consisted of a study coordinator, two field supervisors, eight research assistants, and 10 Neighborhood Health Committee members responsible for facilitating community connections. Community member recruitment used systematic random sampling across four high-density, low-income areas, with low reported uptake of COVID-19 vaccination employing a “spin the bottle” technique to enroll 100 individuals per area, distributed across 10 neighborhoods (10 per neighborhood). Weekend recruitment and data collection strategies were implemented to improve male participation. HCW recruitment followed a convenience sampling approach from the 10 public health facilities, targeting 1-2 professionals from five key departments: HIV/ART, Mother and Child Health, Tuberculosis, Adolescent Health, and Outpatient Services.

For qualitative data collection, the team consisted of four trained and experienced Zambian research assistants overseen by a study coordinator. All sessions took place at six of the 10 health facilities that served a mix of urban, peri-urban, and rural populations. We employed purposive random sampling to enroll a subset of the survey participants, including community members and both professional and lay HCWs, ensuring half of each group was vaccinated against COVID-19 (complete series, with or without booster). Research Assistants contacted selected participants from among those consenting to follow-up during the survey and invited them to participate in an IDI at their preferred time and date or during a scheduled FGD. The RA sent reminders via phone or text message one day prior to the IDI/FGD.

### Ethical Considerations

All participants provided written informed consent in their preferred language (Nyanja, Bemba or English). The study received ethical approval from the National Health Research Ethical Board (AMREF-ESRC P452/2018), Washington University in St. Louis Institutional Review Board (IRB#: 20-09-18), and the University of California, San Francisco Institutional Review Board (IRB#: 23-40096). This study follows the Standards for Reporting Qualitative Research (SRQR) guidelines (**Supplementary File 1)**.^17^

### Data Collection

Both quantitative (survey) and qualitative (IDI and FGD) data collection tools explored individuals’ attitudes, intentions, and beliefs regarding a future adult and adolescent TB vaccine, as well as preferences and recommendations for its delivery.

#### Quantitative Data Collection

The survey was embedded within a broader survey examining COVID-19 vaccine uptake, different disease perceptions and vaccine-specific intentions, childhood vaccine beliefs, and current and trusted vaccine information channels and sources.^16^ Surveys were conducted in community settings, typically at participants’ homes or nearby quiet locations. Prior to beginning the survey, a study team member reviewed disease fact sheets containing standardized descriptions of TB and other conditions, their consequences, and treatment options to ensure uniform understanding across participants (**Supplementary File 2)**. Survey data were collected using the Sawtooth Software Offline Surveys platform.

#### Qualitative Data Collection

Semi-structured guides were used for IDIs and FGDs to explore how participants’ experiences with established vaccines and COVID-19 vaccines might shape their perspectives on a TB vaccine that could be introduced in the near future. The guides incorporated key elements of the ^18^ (BeSD) and 5C (Confidence, Complacency, Constraints, Calculation, Collective Responsibility)^19^ vaccination frameworks. IDIs and FGDs were conducted in quiet, private spaces at the public health facilities serving participants’ communities and were facilitated in participants’ preferred language by experienced Zambian research assistants. All sessions were audio-recorded and lasted on average approximately 120 and 180 minutes for IDIs and FGDs, respectively.

Prior to enrollment, all data collection tools were pilot tested, prompting refinements including shortening the length of the instruments (both the survey and qualitative guides) and modifying language to improve clarity and local language comprehension.

### Data Analysis

#### Quantitative analysis

Descriptive statistics were used to characterize participants and TB vaccine perspectives. Mixed-effects Poisson regression models were used to estimate the marginal probability of participants’ intention to get a new adult TB vaccine upon its availability (defined as “definitely going to get the TB vaccine”); models were adjusted for a minimal adjustment set identified using a directed acyclic graph (DAG)^20^ allowing for random effects at the community (among community members) and healthcare facility (among healthcare workers) levels (**Supplementary Figure 2**). Additionally, we used adjusted mixed-effects Poisson regression models to estimate the marginal probability of participants’ 1) current vaccine information channels and 2) trustworthy vaccine information sources in relation to their TB vaccine intention, with a focus on identifying crucial information sources for vaccine-hesitant individuals. All analyses were conducted using STATA 18.0 (StataCorp LLC, College Station, TX, USA).

#### Qualitative analysis

To facilitate rapid qualitative analysis, immediately following each interview, interviewers completed a structured debrief form, capturing key insights and emblematic quotations from memory and by re-listening to the audio recording. All debrief forms were completed within two days of the respective interview or discussion. Data from all debrief forms were compiled into a data matrix in Microsoft Excel, with each row corresponding to a participant and columns corresponding to a priori key domains of interest systematically captured using the debrief form. The data matrix was analyzed for each stakeholder group (community members, professional HCW, lay HCW) and data type (IDI or FGD) by reviewing each column across all participants or groups. Additionally, each row was examined to ensure interpretations were grounded in the individual’s context. Results were organized according to predefined thematic domains, with interpretation contextualized by participants’ COVID-19 vaccine experiences to understand their influence on TB vaccine acceptance and implementation perspectives and preferences.

#### Triangulation

Following independent analysis of the quantitative and qualitative datasets, we employed a systematic convergence approach to compare findings across methodologies. The principal investigators (ADK and AS) examined quantitative results alongside descriptive qualitative insights to identify areas of alignment, divergence, and complementarity. Specific attention was given to exploring differences between community members and HCWs, comparing their quantitative results with qualitative findings, to develop a comprehensive understanding of perspectives on a new TB vaccine. The primary investigators critically reviewed both corroborating evidence and apparent contradictions, seeking to provide a nuanced interpretation of the integrated findings.

#### Researcher characteristics and reflexivity

The research team included both Zambian and non-Zambian researchers with diverse professional backgrounds that could have influenced data collection, analysis, and interpretation. The team’s own vaccination experiences and professional healthcare roles could have shaped question formulation and interpretation of participants’ TB vaccine perspectives. To mitigate potential bias, the interview guide was developed collaboratively with the Social Science Research Group at CIDRZ, which has extensive local experience with vaccination programs. We partnered with trusted Neighborhood Health Committee members to facilitate community engagement, recognizing that participants might respond differently to community insiders versus outsiders when discussing TB vaccine intentions. All interviewers were fluent in local languages, enabling participants to express nuanced views about TB vaccination in their preferred language. For analysis, ADK took an etic (outsider) analytical approach while AS provided an emic (insider) perspective, with all team members contributing to interpretation. This dual lens was particularly important for understanding both individual and cultural factors influencing TB vaccine intentions. The team regularly discussed potential biases and alternative interpretations throughout the analysis process.

## Results

Overall, 499 participants were enrolled in the survey - 395 (79.2%) community members and 104 (20.8%) HCWs. Across the 25 IDIs and 9 FGDs, 29 community members and 34 HCWs were enrolled. Participant characteristics are summarized in **Supplementary Table 1**.

Below, we present attitudes and beliefs towards vaccines in general and COVID-19 vaccination to contextualize findings on perceptions of and intentions for receiving a new TB vaccine for adults and adolescents, along with recommendations for new TB vaccine delivery.

### Impact of the COVID-19 Pandemic on Attitudes and Beliefs Toward Vaccines

#### Positive Attitude

Generalized trust in vaccines made people receptive to new vaccines, including for COVID-19 and other disease conditions. Both community members and HCWs widely trusted established vaccines like BCG and polio, emphasizing their efficacy as illustrated by the quotes below. Additionally, HCWs observed the pivotal role of vaccines such as polio vaccines in improving public health by reducing disease burden, hospitalizations, and deaths, which bolstered their confidence in the value of vaccination.

*Vaccines are very important to me because when I take a vaccine, I am creating an immune system for that disease to fight against the infection itself.” (professional HCW)*

*”At the end of the day, vaccines will protect us and our children from different kinds of diseases.” (*community member)

Personal experiences with novel adult vaccines strongly reinforced this trust and perceived value of new vaccines, such as those for cholera and COVID-19, as well as those anticipated in the future:

*”…Like with cholera, a lot of people were sick vomiting, diarrhea; like those who got (the vaccine) were fine, including me.” (community members)*

*“…So, I was the one that was advocating that, ‘Let me be the lab rat! If anything goes wrong, you will see it from me…If I survive, you will all go [for COVID-19 vaccination].’ Then the whole household went for vaccines as well because I spearheaded that.” (*community member)

*“This vaccine for COVID…It alerted us on the importance of vaccines…I lost my dad due to COVID. This was a period when we were all refusing to be vaccinated. The clever ones got vaccinated…People were getting very sick and that is when they wanted to come and get vaccinated…But when things became bad, they closed hospitals and all other places…Everything was shuttered, there was no travelling either due to Covid…So if in future, vaccines came, I wouldn’t refuse. I would actually help sensitize more for people to get the vaccines.” (professional HCW)*

#### Variable COVID-19 Vaccine Uptake

Nevertheless, survey data revealed significant differences in COVID-19 vaccine uptake between the two groups, with only 33.4% (132/395) of community members receiving 2+ doses compared to 70.2% (73/104) of HCWs. A substantial proportion of community members (38.0%, 150/395) reported not receiving any doses, compared to just 6.7% (7/104) of HCWs.

#### Skepticism Arising from COVID-19 Vaccination

HCWs offered mixed perspectives on the COVID-19 vaccine based on their professional experiences. While many appreciated the reduction in severe COVID-19 cases following vaccine rollouts, professional HCWs were critical of evolving guidelines around vaccine schedules and booster doses which diminished trust among peers and community members. These experiences, along with breakthrough infections, laid bare expectations about vaccine efficacy and duration of protection for future vaccines. As one lay HCW reflected, *”…If the vaccine is there to protect us, the virus shouldn’t mutate and require another booster.”*

While most participants acknowledged the role of COVID-19 vaccines in mitigating the severity of the pandemic, widespread misinformation and misconceptions created significant barriers to acceptance among others. Many expressed concerns about rapid vaccine development for COVID-19 and insufficient long-term safety data, the absence of which, raised geopolitical concerns, including fears that vaccines might be designed to decimate African populations. One participant noted:

*“…Others were willing and eager to get the vaccine while others were not. Those who were not willing said, ‘It was too early [to have a vaccine for COVID], maybe they want to wipe us out as Africans…All these years we have been [unsuccessfully] crying for an HIV vaccine, but how come the COVID vaccine is here? Maybe it is part of the propaganda’…” (lay HCW)*

Concerns about rapid vaccine development and falsely attributed side effects shared below created enduring skepticism about newer vaccine technologies as voiced by one HCW:

*”…Due to the reactions that come up after someone is vaccinated, I think COVID-19 vaccines have brought more issues for other future vaccines for people to be accepting them…We have even seen people claiming that others have died because of taking a vaccine without proper facts and evidence.” (lay HCW)*

Side effects were a prominent concern among participants and social media amplified rumors about infertility in women, low libido in men, and spurious claims about magnets sticking to vaccinated bodies. As one HCW observed:

*“When COVID came, we were all scared of getting vaccinated because people said a magnet would hold [stick to skin], or you will have certain side effects. I said I won’t get it, but with time I realized that it was a worst of time, so I got vaccinated.” (professional HCW)*

They later went on to say,

*”People are skeptical. People were refusing, saying that they heard stories that people will have magnets attaching to their bodies. They were just rigid, no matter how much we would educate them…” (professional HCW)*

Both professional and lay HCWs acknowledged that while the pandemic heightened public awareness of the importance of vaccines, it also underscored the need for transparent, consistent communication to sustain trust when introducing new vaccines. As one community member noted, *”people will always be people”* and will be skeptical about something they don’t know.

#### Accessibility challenges for COVID-19 vaccination

Commonly mentioned accessibility challenges included long travel distances to vaccination sites, transportation costs, inconvenient service hours, and vaccine stockouts. One community member shared their difficult journey in getting a COVID-19 vaccine:

*“Where am I going to get the vaccine from? Like the first time I wanted to get the COVID vaccine, I came to this same facility A. They never had the vaccine here…I went to facility B; I stood for some hours and then I was just told, ‘No, we are not giving the vaccine here. Try some other day.’ Until after doing some research online I found that at Facility C is where most people were getting their COVID jabs…So, I had to travel all the way that side, probably that 8-10 kilometers distance…that gives it about 30 kilometers of traveling [to and from the three facilities]…“ (*community member)

Additionally, time constraints, especially for individuals with erratic work schedules, posed obstacles to getting vaccinated. Gender dynamics created additional barriers - some women required spousal permission for vaccination, particularly when visiting facilities for other reasons, while some men refused to allow their wives or children to be vaccinated altogether. Religious beliefs and fatalistic attitudes, such as trusting only in “*God’s time*” and ancestral spirits such as those among the “*Ba Zezuru,*”^21^ transferred to COVID-19 vaccine hesitancy despite targeted education efforts. Further, some churches refused to allow vaccination within their premises.

HCWs reported multiple logistical challenges that complicated vaccine delivery and reduced access, including insufficient transportation resources, delayed vaccine delivery from the Ministry of Health (MOH) to sites, and health workers’ lack of motivation due to inadequate financial support. Language barriers in areas where HCWs were not fluent in local dialects created additional communication challenges. Additionally, HCWs said that community members often demanded to see HCWs vaccinated first to trust COVID-19 vaccine safety, while some resorted to strategies like alcohol consumption to disqualify themselves from vaccination.

### Perceptions and intentions for a new TB vaccine

#### TB Vaccine Prioritization

TB emerged as a high priority for new vaccine development, ranking second and third (out of 7 disease-specific vaccine candidates) among healthcare and community members, respectively (**Figure 1**). 71.1% of HCWs and 58.7% of community members placed TB vaccine development in their top three priorities, perceiving TB as a major community problem being airborne, having high transmissibility in public spaces, and being deadly. Both community members and HCWs highlighted the limitations of existing TB preventive measures, including TB preventive treatment (TPT), underscoring that improvements in ventilation and cough etiquette are insufficient on their own to fully control the spread of TB.

**Figure 1.**
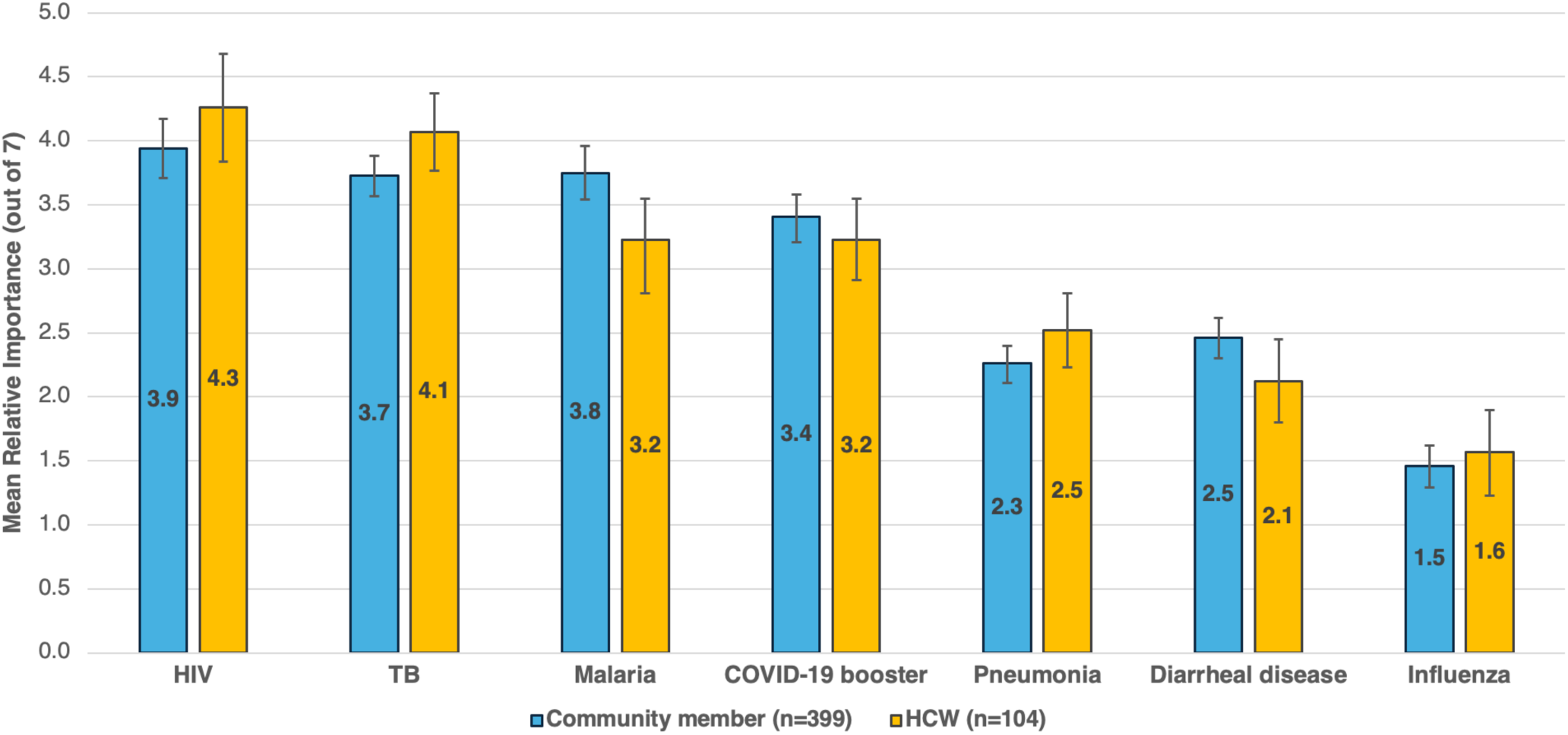
Mean relative importance of disease-specific vaccines to be prioritized for development and introduction according to community members and HCWs (N=499).

*“…So that we stop talking about how many people are dying from TB – yes, the treatment is there but do we have something to prevent? Prevention is better than cure because prevention is cheap compared to treatment…People are dying…So, if there is capacity to develop such vaccines, better go ahead…” (professional HCW)*

*“People in the community are concerned about TB because it kills faster than HIV and they are also concerned about taking tablets for prevention [TPT]. Instead, they would prefer an injection that they could get once…which is better.” (lay HCW)*

When considering the potential benefits of TB vaccination, community members placed emphasis on personal and social dynamics, such as the fear of ostracization if they were to get TB and the impact of TB on daily life and ability to work. HCWs focused on their frequent exposure to undiagnosed patients and the complexities of managing long and toxic TB treatment regimens.

#### Intention to Get a TB Vaccine

The prioritization of a new TB vaccine was matched by high intention to vaccinate, with 77% of community members and 83% of HCWs indicating they would definitely get a TB vaccine if recommended. Perceived TB risk (11.4% very unlikely to get TB to 58.3% very likely to get TB) and concerns about severe outcomes, including death (4.2% not at all concerned to 71.9% very concerned), if they developed TB, were the only factors that significantly drove TB vaccine intention (**Figure 2, Supplementary Table 2)**. Among HCWs, 73.1% believed they were “very likely” to develop TB compared to 54.4% of community members, while 79.8% of HCWs and 69.9% of community members expressed being “very concerned” about TB’s severe consequences, including death if untreated (**Figure 3**). This aligns with participants’ reports that they got the COVID-19 vaccine as COVID-19 deaths increased, recognizing its benefits due to fewer side effects and milder disease in those vaccinated. Nonetheless, previous COVID-19 vaccine uptake showed no significant association with intention to receive a future TB vaccine (**Figure 2, Supplementary Table 2)**.

**Figure 2.**
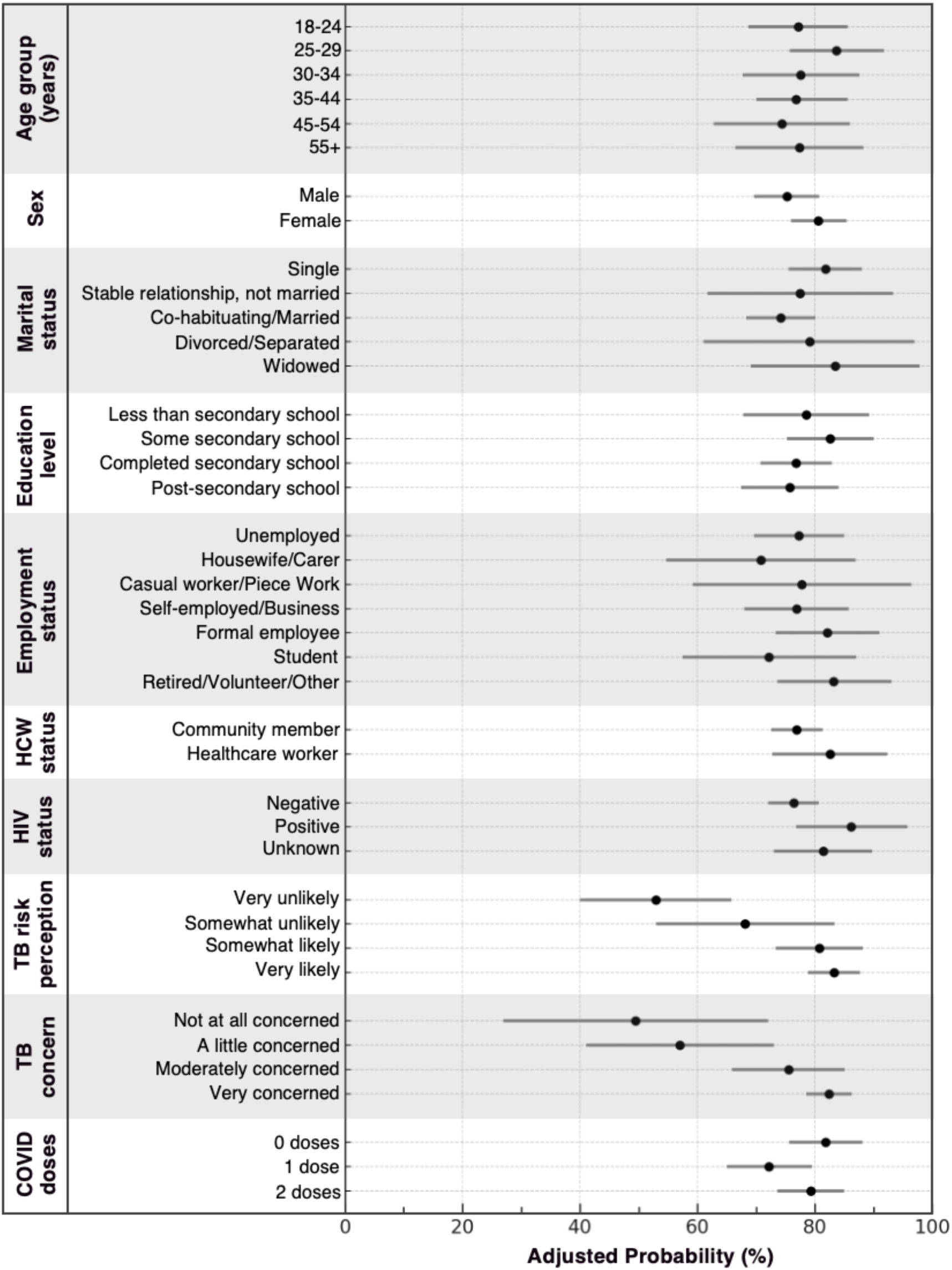
Predicted probability of intention to get a future TB vaccine by subgroup.

**Figure 3.**
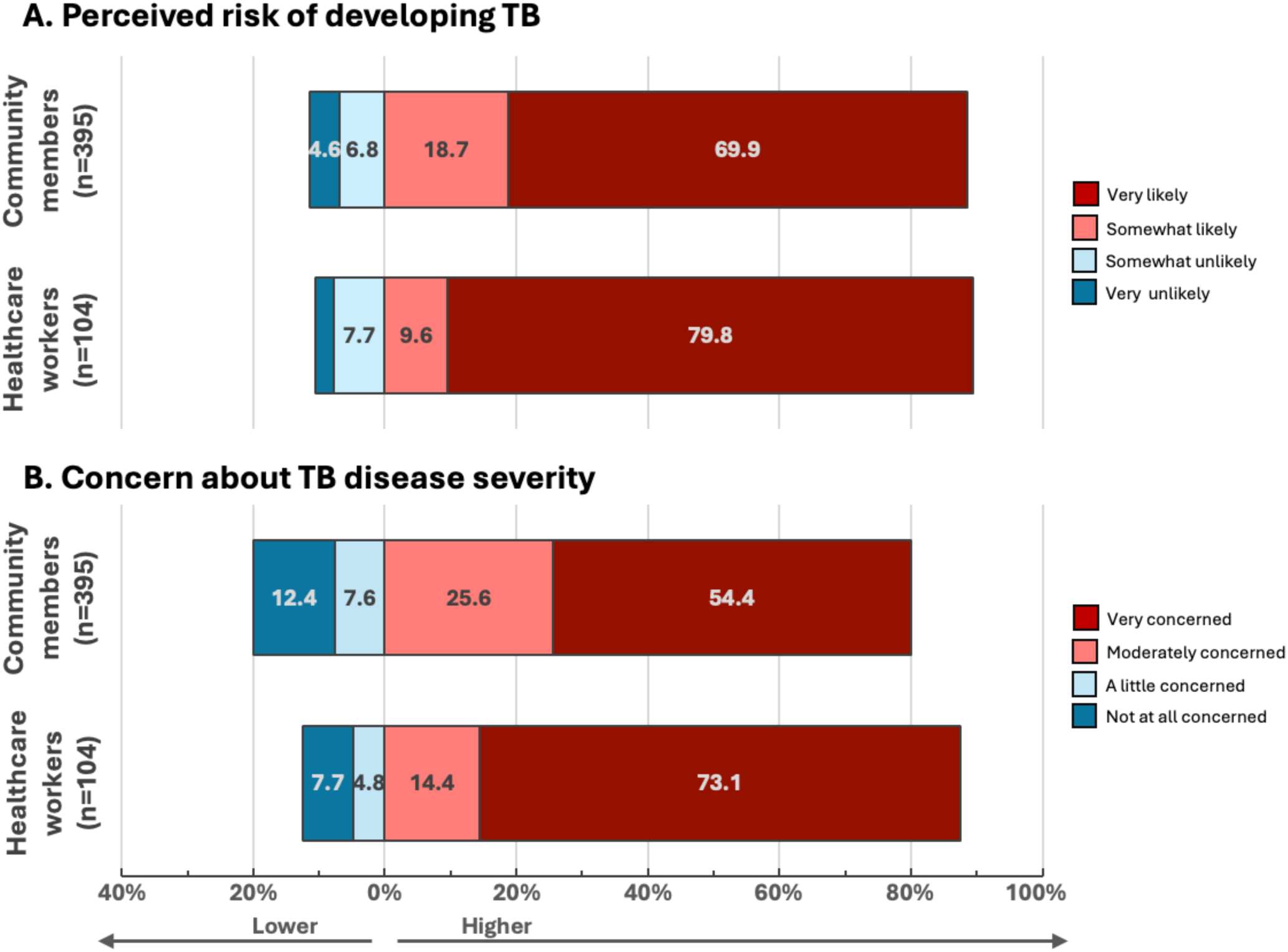
TB Risk and Severity Perception among Community Members and HCWs.

#### Acceptable TB Vaccine Attributes

Though opinions on acceptable and ideal TB vaccine attributes varied, nearly all participants demonstrated a pragmatic willingness to accept TB vaccines that met most of WHO’s minimum requirements^22^ (See **Table 1)**.

**Table 1.**
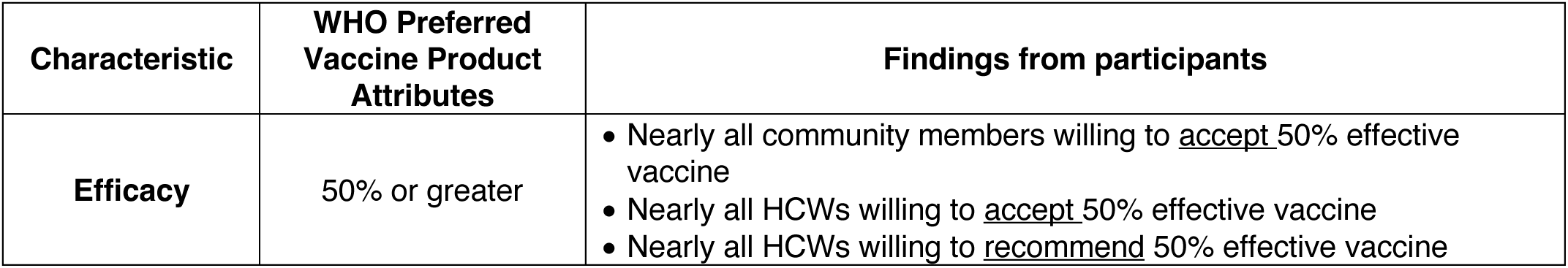
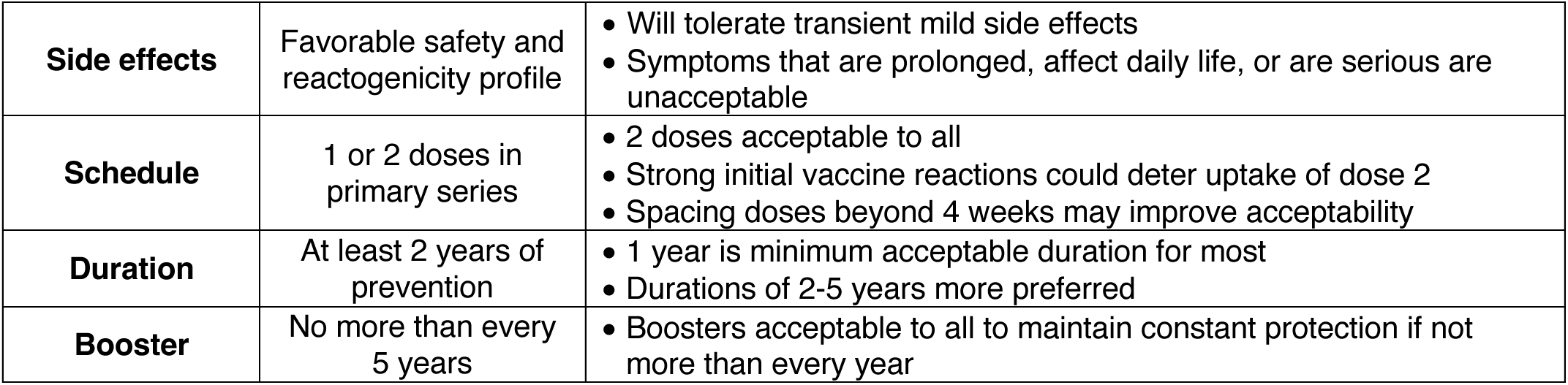
Participant Preferences for a New TB Vaccine in Relation to WHO Preferred Vaccine Product Attributes ^22^.

| Characteristic | WHO Preferred Vaccine Product Attributes | Findings from participants |
| --- | --- | --- |
| Efficacy | 50% or greater | Nearly all community members willing to <u>accept</u> 50% effective vaccine<br>Nearly all HCWs willing to <u>accept</u> 50% effective vaccine<br>Nearly all HCWs willing to <u>recommend</u> 50% effective vaccine |
| **Side effects** | Favorable safety and reactogenicity profile | • Will tolerate transient mild side effects<br>• Symptoms that are prolonged, affect daily life, or are serious are unacceptable |
| **Schedule** | 1 or 2 doses in primary series | • 2 doses acceptable to all<br>• Strong initial vaccine reactions could deter uptake of dose 2<br>• Spacing doses beyond 4 weeks may improve acceptability |
| **Duration** | At least 2 years of prevention | • 1 year is minimum acceptable duration for most<br>• Durations of 2-5 years more preferred |
| **Booster** | No more than every 5 years | • Boosters acceptable to all to maintain constant protection if not more than every year |

##### Efficacy

Views on acceptable vaccine attributes revealed nuanced preferences, particularly regarding efficacy. A wide range of efficacy levels ranging from 40% to 99% were cited, however, 50% efficacy threshold was deemed acceptable by nearly all community members and HCWs. HCWs consistently expressed preference for higher efficacy levels, with professional HCWs frequently citing a threshold of ≥80% due to their occupational exposure. One professional HCW shared that, *“If its 50/50, the fighting power would be neutral…I would get [vaccinated] but it [vaccine efficacy] should at least be more [than 50%].”* Some HCWs specifically rejected a 50% efficacy level, with one noting that current TB prevention methods [TPT] already offered better protection. However, most participants emphasized that partial protection was better than none, as one participant remarked, “*If it protects at least a little, it’s better than nothing*.”

##### Side effects and safety

As with the COVID-19 vaccine, participants expressed concerns about potential side effects from a new TB vaccine, specifically headache and gastrointestinal issues (vomiting, diarrhea), followed by loss of appetite and rashes. While mild, short-term effects were considered tolerable, participants would reject a vaccine with prolonged (greater than several days), debilitating symptoms affecting daily activities, particularly those involving the brain or heart. One participant noted:

*“As long as the vaccine makes one to feel sick - [That] is what makes people to stay away from taking a vaccine because a vaccine is supposed to be protecting me but instead it is making me feel sick in the process…So, side effects can be there, but they should not be severe…”*

Reminiscent of COVID-19 times, some participants also sought information about safety during pregnancy.

##### Schedule, duration, and booster

All participants indicated they would accept a two-dose TB vaccine schedule, emphasizing that completing the full course was essential for achieving full protection, with one lay HCW emphasizing “*health is wealth*.” Community members and lay HCWs specifically noted that proper explanations and education about the dosing schedule would enhance acceptance with one respondent anticipating that based on COVID-19 roll-out, community members would ask questions like, *“If you give me this vaccine, am I supposed to come and get another one?”* One HCW anticipated feeling reluctant but accepting a second dose like they did during COVID-19. A community member who though accepting, said they would prefer at least 6 months between doses, expressing concern that a 4-week second dose interval, as used with COVID-19 vaccines and current TB vaccines in development, was too short.

Regarding duration of protection, most participants indicated that one year was the minimum acceptable period, recognizing it as sufficient to provide ongoing protection and reduce doubts about the vaccine’s efficacy. Many expressed a strong preference for longer durations, with one participant expressing a willingness to wait for a vaccine providing five years of protection, considering it optimal for long-term disease prevention. All participants were willing to accept boosters provided the interval between doses was at least one year, to achieve sustained protection, with some describing it as necessary once the vaccine “*expires*” or “*loses power*.” One community member noted that requiring boosters any sooner than a year could lead to doubts about the vaccine’s effectiveness.

#### Perceived Benefits of a TB Vaccine

##### Protection for self and loved ones

Community members often framed their willingness to vaccinate using strong metaphors, likening the vaccine to “*bulletproof*” protection or “*putting on a combat*,” highlighting their belief in its protective role. Many participants (both community members and HCWs) noted that protecting oneself served as a primary motivator for getting vaccinated, driven by the high-risk perception and concern about the severity of TB described above (**Figure 3**). Most participants also viewed vaccination as a crucial way to protect their families and close contacts.

##### Protection against occupational exposures

HCWs expressed particularly strong motivations due to their unavoidable occupational exposure reinforced by their COVID-19 experiences and the burden of lengthy treatment:

*“…We just [know] them as normal patients, not knowing the cough they have is TB…Some even cough on us.” (Professional HCW)*

*”The health care workers would be very happy [to get a TB vaccine] because we are at high risk. You know even with me, I got COVID 3 times that time. So, if the vaccine came, we would get it with my friends.” (Professional HCW)*

*”I have seen those who take drugs for 6 months, and sometimes they increase to 8 months, so I see them struggle…I am first in being scared of medication even though I educate people about taking medications, even injections. If you brought one here, we would need to negotiate.” (Professional HCW)*

Beyond healthcare settings, participants identified miners, prisoners, and market vendors as groups who would particularly benefit from a TB vaccine due to their exposure in crowded or poorly ventilated environments. Those working in industrial settings were also highlighted as potential beneficiaries.

##### Protection against biological, behavioral, and social risk factors

Participants identified several additional groups as particularly vulnerable to TB and who would benefit from a vaccine. PWH were highlighted due to compromised immunity, while children were emphasized despite prior BCG vaccination due to their developing immune systems. Other vulnerable groups mentioned included pregnant women, the elderly, and those with conditions like diabetes.

Participants highlighted how certain behaviors and social patterns increased TB risk and emphasized the potential role of vaccination in mitigating these risks. High-risk behaviors, particularly alcohol consumption, were noted as increasing vulnerability with one participant observing that

”…*A lot of people that we see in the community have coughs, especially those that drink kachasu [local spirit]…They have a lot of TB so it would be good to bring the vaccine to reduce the spread of the disease.” (community member)*

The airborne nature of TB and challenges of avoiding exposure in daily life, for example in crowded buses, markets, and schools, further heightened the perceived value of vaccination with one community member noting that, *”TB is airborne, so if you are vaccinated you can socialize with friends without problems and getting sick; it [vaccination] is very necessary.”* Adolescents and young adults were seen as particularly vulnerable due to their social mixing patterns and environmental exposures.

##### Broader societal benefits

Beyond individual protection, participants emphasized how vaccination could benefit society more broadly. HCWs saw vaccination as an opportunity to both prevent disease and address misconceptions about TB. As one HCW stated, “*We need to break the myths that TB cannot be cured. Education and vaccination could really help us manage this better.”* HCWs also valued maintaining their ability to care for patients, recognizing that getting TB would impair their work capabilities. Lay HCWs noted that vaccination could improve patient connections by eliminating the need for stigmatizing masks that create barriers with patients.

#### Preparing for and Implementing a New TB Vaccine

Participants provided comprehensive recommendations for TB vaccine delivery and communication strategies tailored to different target populations. Key implementation strategy features identified for adults, adolescents, and healthcare workers are summarized in **Table 2** and described further below.

**Table 2.** Key recommended features of delivery and communication strategies for a new TB vaccine among adults, adolescents, and healthcare workers.

| Target Recipient | Delivery Strategies | Communication Strategies |
| --- | --- | --- |
| **Adults** | • **Facility-Based Delivery:** Integration into routine visits; trusted sites<br>• **Community-Based Delivery:** Use of convenient venues: churches, markets, workplaces<br>• **Door-to-Door Campaigns:** Reaching the elderly, mobility limited, rural settings with limited access<br>• **Mass Campaigns:** Weekend hotspots, football fields<br>• **Gender-Specific:** Women at markets/salons, men at bars/workplaces<br>• **Practical Support:** Transport allowances, consistent supply, flexible timing<br>• **Trust Building:** Witnessed vaccination events, especially HCW vaccination | • **Sensitization:** early and ongoing to dispel myths and build trust<br>• **Trusted Sources:** MOH, HCWs, community and religious leaders<br>• **Accessible Channels:** TV, health facilities, radio, social media<br>• **Engaging Methods:** Use of drama, storytelling, and testimonials.<br>• **Group-Specific:** Local language materials, visual aids<br>• **Community Engagement:** Dedicated vaccine communication groups |
| **Adolescents** | • **School-Based Delivery:** Primary setting, leveraging HPV/COVID experience<br>• **Community-Based Delivery:** Sports events, churches<br>• **Door-to-Door Campaigns:** Reaching non-school youth<br>• **Mass Campaigns:** With engaging activities/games<br>• **Youth-Specific:** Youth corners, recreation centers<br>• **Practical Support:** Parent consent processes, flexible scheduling<br>• **Trust Building:** Peer role models, vaccinated adolescents | • **Trusted Sources:** Parents, teachers, peers, HCWs<br>• **Accessible Channels:** Schools, social media, interactive online platforms<br>• **Engaging Methods:** Use of drama, interactive discussions, sports events<br>• **Group-Specific:** Age-appropriate and relatable content, simple language<br>• **Parent Focus:** Targeted education for buy-in<br>• **Community Engagement:** Youth groups, school programs |
| **HCWs** | • **On-Site Delivery:** Vaccination at health facilities, ideally without needing to leave post<br>• **Flexible Scheduling:** Availability during different shifts and/or off-hours<br>• **Integrated Approaches:** Offering vaccines alongside routine HCW check-ups or trainings<br>• **Department-Specific Programs:** Targeting key units (HIV/ART, MCH, TB, Adolescent Health, OPD)<br>• **Trust Building:** Use of peer influence and senior staff modeling acceptance<br>• **Government Mandates:** Clear policy directives and requirements | • **Trusted Sources:** Messaging from senior clinicians, MOH representatives, and respected peers<br>• **Workplace Channels:** Use of facility notice boards, WhatsApp groups, and staff meetings for updates<br>• **Professional Channels:** WHO endorsements, medical journals, and conferences<br>• **Evidence-Based Information:** Comprehensive safety, efficacy, and duration data with transparent development and manufacturing processes<br>• **Dialog-Based:** Opportunities to ask questions or discuss concerns in group forums or during ward rounds<br>• **Testimonial Sharing:** Peer stories of positive experiences with vaccination |

#### Early Preparation and Sensitization

##### Early community engagement

Participants emphasized beginning preparation and communication at least 3-6 months before vaccine availability. Both groups stressed how early, continuous education, and thorough community engagement could help build vaccine confidence, address misconceptions before they take root, and ensure participation as illustrated by the quotes below.

*”When the healthcare workers are going round in the community, we don’t know why they are injecting us. Of course, we know there is cholera, TB, and other diseases, but if I don’t have information, I will refuse because I do not want to get into something that I don’t know because the government has said that they should give me an injection. This is my life and this is my health, this is not the governments’ body…So, they need to go into the community and engage stakeholders who can sensitize.” (Community member)*

*”…They pass through today and tell us that tomorrow you should come to such and such a place to get an injection…They tell us at gunpoint. They do not tell us even one week before because you know that some people work and cannot manage to go and get an injection the same day…” (Community member)*

*”What is important is education about the same vaccinations, you need total sensitization; people are difficult. Use media such as megaphone and radios, tell people the importance of vaccines - People can’t just accept to be vaccinated, education is number one.” (Professional HCW)*

Early sensitization is also needed to address specific cultural barriers. While the potential for stigma associated with a TB vaccine was overall felt to be minor, some noted an association between TB and HIV that could deter vaccine uptake. Campaigns should normalize TB vaccines by making it clear that TB vaccines are for everyone, while also addressing religious opposition to vaccination such as among the “*Ba Zezuru*” as described above.

##### Vaccine information channels and trusted sources

Survey data revealed that television was the most common channel for receiving vaccine information (80.6%), followed by health facilities (67.3%), social media (60.5%), radio (57.1%), word of mouth (53.3%), and church (50.3%), with information sources differed between community members and HCWs (**Supplementary Table 3**). After adjustment, there were few differences in vaccine information channels by TB vaccine intention status, although there was a trend toward word of mouth and social media being slightly more common channels among those with lower intentions to get a TB vaccine, and churches being a less common channel (**Supplementary Table 4**). In IDIs, participants recommended diverse communication approaches, including mass media, door-to-door outreach, and community gatherings. They underscored the importance of consistent and repeated communication to sustain awareness. Creative methods such as drama performances, storytelling, and testimonials were suggested to make information more accessible and compelling.

Survey findings identified the MOH (87.6%), HCWs (69.5%), and Government (55.7%) as the most trusted sources for vaccine information; religious leaders (18.8%), scientists (18.6%), community leaders (13.4%), friends and family (12.4%) and social media (9.0%) were less trusted overall, with community members generally reporting lower trust across sources compared to HCWs (**Supplementary Table 3**). Trusted sources showed few differences by TB vaccination intention after adjustment, except for those less willing to vaccinate who had slightly lower trust in community leaders and slightly higher trust in social media (**Supplementary Table 4**). However, qualitative data highlighted how personal relationships with diverse community figures, including religious leaders and herbalists and significant others could reduce resistance and strengthen community buy-in. As one community member explained*, “…significant other telling me - that is the person I trust. Yes,…I can go get it.”*

Some participants suggested forming dedicated vaccine communication groups to serve as community educators and messengers. These groups would travel for community gatherings, explain vaccine mechanisms and disease information, and engage in public education - though with an emphasis on supportive rather than intimidating approaches. This could provide a structured bridge between formal communication channels and community-level engagement.

##### Tailored Approaches for Different Audiences

HCWs emphasized adapting communication channels and messaging for different audiences. Men were best reached through workplace communications and community leaders, while women were more receptive to health facility-based information and community health worker outreach. For elderly individuals, simple messaging with visual aids and local language materials was essential.

For adolescents, communication needed to be age-appropriate and engaging. While schools provided a structured channel through teachers, social media platforms were crucial for reaching this tech-savvy group. Interactive approaches like drama and peer education were recommended to increase relatability. As one lay HCW noted: *”The adolescents, the way they are, as long as it is their fellow adolescent that is handling them, they are okay.”*

##### Key Information Content

Community members emphasized needing comprehensive information to make informed decisions about new vaccines. They stressed the importance of understanding potential side effects, vaccine effectiveness, dosage requirements, and the mechanisms of action. Clear and thorough counseling from trusted HCWs was deemed essential to set accurate expectations and provide clarity on the vaccine’s benefits and risks. As two community participants noted:

*”I should be told what side effects each vaccine has, because they say that these are the side effects and therefore do not be surprised.” (Community member)*

*”…Telling them about how the vaccine works, that it prevents TB and telling them that if you are vaccinated, you won’t get sick.”(Community member)*

A remnant of COVID-19 vaccine skepticism, participants stressed the importance of transparent communication about the vaccine’s origin and manufacturing processes to build credibility with many preferring to wait and observe the experiences of early vaccine recipients to determine vaccine safety and confidence.

#### Adult Delivery Strategies

Building on COVID-19 experiences, participants recommended a blend of facility-based and community-driven vaccination efforts. While health facilities were trusted sites, many noted accessibility barriers. Community venues such as churches, markets, and workplaces were highlighted as effective alternatives due to their convenience and familiarity. Reflecting on the success of similar strategies during COVID-19, one community member described how, on *’COVID [vaccination] days*,’ HCWs went to schools, companies, and churches, making vaccines more accessible, which helped dispel myths, build trust, and encourage uptake.

*”…I think what worked more was following people in the communities and not them coming to the facility, no. So, following people in the community was much easier for the fact that COVID-19 had a lot of myths…They know to say if you are in the market, you are in the market. It is even cheaper for them; they are not going to spend any money on transport…that one worked really well.” (Professional HCW)*

HCWs echoed the importance of community-focused strategies, advocating for mobile clinics paired with education initiatives to promote vaccine acceptance. They suggested that HCWs should be first to receive vaccines to demonstrate safety, addressing a noted gap from COVID-19 implementation. One community member agreed, noting, “*You can’t trust something you have not seen*.” Gender-specific considerations were also felt to be important, with recommendations for targeting women at markets, homes, and salons, while reaching men through community spaces and bars (with protocols for intoxication). Mass vaccination campaigns were recommended, particularly at weekend hotspots like football fields with advanced community sensitization to ensure turnout. Door-to-door campaigns were emphasized as essential for reaching those with limited mobility or in rural areas, provided familiar volunteers delivered messages and administered vaccinations.

Practical considerations mentioned included ensuring consistent vaccine supply, maintaining cold chain logistics, and tailoring delivery times to accommodate different populations. Some suggested offering additional health services, like blood pressure checks, as well as incentives for vaccine uptake and as for COVID, even vaccine requirements to attend specific venues and events.

*“Like it happened during Covid-19, they just said no one will enter the gate if not vaccinated. So we all went to get it, we took ourselves.” (Community member)*

#### Adolescent Delivery Strategies

Schools were consistently identified as the primary setting for adolescent vaccination, offering a centralized location and existing infrastructure for reaching this population, with HPV and COVID-19 vaccination programs cited as successful models. Both community members and HCWs emphasized the crucial role of teachers in facilitating parental consent and promoting vaccine uptake. As one participant noted:

*“It is best to use schools because that is where they [adolescents] are found. Go through the teachers who will tell them about why you are there” (Community member)*

Alternative venues included sports events, churches, and door-to-door campaigns for reaching non-school-going youth. Though suggested as complementary venues, some questioned the effectiveness of delivering vaccination at youth-friendly spaces within healthcare facilities due to low utilization.

Family influence emerged as a critical factor in adolescent vaccination decisions. Participants recommended conducting parallel education campaigns for parents and adolescents. Mass vaccination campaigns were mentioned as promising if paired with engaging activities and games. Peer-led initiatives, with vaccinated adolescents serving as role models, were also recommended to foster trust and relatability. As one HCW noted:

*“If they [adolescents] find an adult and as long as they know that they may be known by the adult, then that will be a problem…They will be thinking that their mothers at home will know that they got a TB vaccine. But if it is their fellow young person [that is handling them], they have no problem with that.” (Lay HCW)*

#### HCW Vaccination and Delivery

##### Factors Influencing HCW Vaccine Acceptance

HCWs emphasized needing comprehensive, evidence-based information about vaccine safety, efficacy, and duration of protection before accepting vaccination themselves because, *“I can be a health care worker but maybe I don’t have information on that vaccine that they have brought…so information would be really needed.”* They advocated for transparency about vaccine development, including trial results and testing processes, and noted the strong influence of WHO Endorsements and of government mandates, with one HCW noting that: *”If the government says it is a requirement, most healthcare workers would participate.”* Additionally, senior healthcare professionals modelling vaccine acceptance would demonstrate vaccine safety to both staff and community members.

##### Supporting HCWs in Vaccine Delivery

For effective vaccine promotion and delivery to communities, HCWs emphasized needing practical support including transport allowances, flexible scheduling, and adequate staffing. Professional recognition, peer support networks, and monetary incentives were identified as important for maintaining motivation throughout vaccination campaigns, with one HCW noting that, *”if they attach something at the end of the day to encourage the healthcare worker, something monetary.”*

## Discussion

This mixed-methods study in Lusaka, Zambia reveals high baseline acceptance and pragmatic flexibility regarding characteristics of a future adult TB vaccine among community members and HCWs. Our findings suggest that despite challenges experienced during COVID-19 vaccine rollouts and limited COVID-19 vaccine uptake, both community members and HCWs highly prioritized TB for vaccine development and introduction, driven by concerns over its airborne transmission, serious health consequences, and the limitations of current preventive measures. Perceived susceptibility and severity created a strong intent to receive a TB vaccine in both groups. Participants accepted the minimum thresholds for all WHO’s preferred vaccine attributes,^22^ though HCWs showed a preference for higher efficacy thresholds given the comparative advantage of already available TPT regimens.^23^ Participants widely accepted a two-dose vaccine schedule, provided that adequate education and spacing between doses were ensured. Successful implementation will depend on early and sustained engagement, trusted messengers, targeted delivery approaches, and policy alignment with community needs and preferences.^14^

The high reported intention to accept a future TB vaccine (77% community members, 83% HCWs) is particularly noteworthy given this study setting’s historically low COVID-19 vaccine uptake. These findings align with a study of pregnant women in Ethiopia that found a high willingness (>90%) to receive a future TB vaccine while pregnant or lactating despite low acceptance of COVD-19 vaccination.^24^ This suggests that TB-specific factors, such as perceived risk, disease severity,^25,26^ and limitations of current prevention methods, may override general vaccine hesitancy. However, these acceptance rates must be interpreted cautiously. Past experiences with other vaccines, including COVID-19, underscore the gap often observed between stated intentions and real-world uptake once questions of logistics, trust, and messaging come into play.^27^ While our findings highlight strong baseline intentions, actual demand could be undermined if challenges related to service delivery, misinformation, or changing guidelines persist.

Participants’ perspectives on acceptable vaccine attributes have important implications for both vaccine development, adoption, and implementation. The willingness of both community members and HCWs to accept vaccines meeting minimum WHO specifications (50% efficacy, two-dose schedule)^14^ suggests that even partially effective TB vaccines could achieve meaningful uptake if clearly communicated and delivered through accessible channels.^6,8,10,28^ Nonetheless, the marked preference for higher efficacy, particularly among HCWs, indicates that public health messaging must transparently address any efficacy gaps and emphasize the collective and individual benefits of even partial protection while managing expectations about vaccine performance.

Implementation strategies identified in our findings suggest that the successful rollout of a new TB vaccine will require mixed facility-based and community-driven approaches, including door-to-door outreach, mass weekend vaccination events, and school-based programs for adolescents.^5,6,12,13,28^ Gender-specific strategies emerged as particularly important, such as targeting men through vaccination events at workplaces and bars and women at markets.

COVID-19 pandemic experiences were central to participants’ perspectives on a TB vaccine. Many respondents described how misinformation about side effects, shifting guidelines on booster doses, and inconsistent service delivery eroded trust. Conversely, these experiences underscored that early, targeted communication campaigns addressing misunderstandings head-on can improve uptake. Together, these insights suggest that the potential success of a TB vaccine rollout depends on anticipating and proactively addressing concerns about safety, dosage schedules, and side effects.

Participants emphasized that these efforts should begin at least 3-6 months before vaccine availability to build confidence and address misconceptions before they take root. This means ensuring consistent guidelines, avoiding abrupt changes in dosing recommendations, and engaging trusted community messengers early. The approach requires proactive education, transparent communication, phased implementation, and leveraging social proof through early adopters such as HCWs.

Participants identified HCWs as a unique bridge between public health authorities and communities. Because of their own high-risk exposure to TB, HCWs stand to benefit directly from vaccination, and they can also serve as the most trusted voices to advocate for its acceptance. However, experiences from COVID-19 revealed that HCWs themselves need clear, evidence-based information on safety, efficacy, and the vaccine’s development process to confidently adopt and champion it. Moreover, meaningful support such as transport allowances, increased staffing, and flexibility in scheduling community outreach emerged as critical for sustaining their motivation and capacity to vaccinate broader populations.

Beyond HCWs, participants also highlighted the importance of diverse communication channels and information sources. Television, radio, and health facilities emerged as dominant sources of health information. While the MOH and HCWs maintained the highest trust levels across all groups regardless of TB vaccine intentions, a similar proportion of participants reported religious leaders as a trusted source as scientists and vaccine developers, and word-of-mouth from trusted friends and family was both commonly used and influential among participants, particularly those with lower vaccine intentions. This necessitates layered communication strategies leveraging both mass media and interpersonal channels while capitalizing on the consistently high trust in official health authorities.

While the potential for stigma associated with a TB vaccine was overall felt to be minor, the association between TB and HIV-related stigma, as well as some fears of ostracization, suggests that targeted interventions will likely be needed to normalize TB vaccination.^29,30^ Integrating the TB vaccine into routine immunization schedules and public health initiatives can help reduce any stigma and encourage widespread acceptance.^7,12^

Several important implementation considerations emerged that could help ensure that the high stated acceptance of a new TB vaccine translates into robust uptake and warrant attention from policymakers (**Supplementary Table 5**).^31^ These include establishing clear communication protocols, ensuring adequate HCW training and support, developing flexible delivery mechanisms, creating community engagement frameworks, and implementing monitoring systems for vaccine confidence and uptake. Drawing from Diffusion of Innovations theory, the Bass model offers a useful framework for sequencing these strategies across adopter groups. While reported willingness to vaccinate was high, the model remains relevant in bridging the gap between intention and actual uptake. The Bass model distinguishes between external influences (e.g., mass communication, official recommendations) and internal influences (e.g., peer networks, social proof). Most HCWs and some high-risk community members are likely to be early adopters who respond primarily to external influences and transparent, evidence-based information. In contrast, broader community uptake may depend more on internal influences, such as observing and hearing about positive experiences from vaccinated peers and trusted individuals. By applying lessons from COVID-19 and focusing on culturally appropriate, community-driven strategies, public health programs can mitigate vaccine hesitancy and ensure new TB vaccines reach those at greatest risk.

Our study had several methodological strengths, including the inclusion of community members and healthcare workers (both professional and lay) – two crucial stakeholder groups for successful TB vaccine rollout –, systematic random sampling from households across multiple communities with historically low COVID-19 vaccine coverage, and robust triangulation of quantitative and qualitative data sources using well-established vaccination frameworks.^18,19^ However, our findings should be interpreted within several limitations. The study’s focus on urban/peri-urban Lusaka may limit generalizability to rural areas where access challenges and community dynamics differ. The selection of areas with low COVID-19 vaccine coverage may have biased results, though this provided valuable insights into intentions and barriers among more hesitant populations. The convenience sampling of HCWs could introduce selection bias, potentially overrepresenting those more engaged with vaccination programs. Social desirability bias may have inflated reported vaccine acceptance rates, particularly in face-to-face interviews, leading to overestimation of actual uptake intentions.^27^ Future research should examine perspectives in rural areas and other regions of Zambia to inform national implementation planning. Studies exploring specific communication strategies, including specific messages, and their effectiveness would be valuable for refining engagement approaches.

In conclusion, while challenges lie ahead, our findings among community members and HCWs suggest strong potential for successful rollout of a new TB vaccine in Zambia, provided that policymakers and implementers heed the lessons from COVID-19. Early and comprehensive preparation, incorporating the specific recommendations identified in this study, including robust HCW support, transparent, consistent communication, and person-centered delivery will be critical for a successful TB vaccine rollout that could prevent millions of new TB cases and deaths in high burden settings like Zambia

## Acknowledgments

We would like to thank all participants for their time as well as the healthcare facilities that provided their support for this study. Claude (Anthropic) was used for copyediting and language refinement during manuscript preparation. All AI outputs were reviewed and approved by the authors before inclusion.

## Contributions

ADK and AS conceptualized and designed the study. ADK, NL, JMP, MH, KS, SSS, EHG, RML, and AS designed the data collection tools. MH, BS, NZ, HN, EKN coordinated and implemented data collection activities. ADK, JMP, AS wrote the statistical analysis plan. ADK, JMP, and AS cleaned and analyzed the data. ADK and AS drafted the manuscript with substantial input from all authors. All authors reviewed the manuscript and were responsible for the decision to submit. NL, JMP, and ADK had access to and verified all the data.

## Data Availability

A de-identified quantitative dataset supporting the conclusions of this article will be made publicly available in the Dryad Digital Repository upon publication of this manuscript. Qualitative data will be shared upon reasonable request to ensure authors are involved in context-driven interpretation.

## Patient and Public Involvement

Patients and the public were not involved in conducting this study. However, local health professionals, researchers, and organizations were consulted in designing and carrying out study activities and interpreting results.

