## Supplementary Materials for "“Prevention is better than cure” - A Mixed Methods Study of Communities’ and Healthcare Workers’ Perspectives on New Adult and Adolescent TB Vaccines and their Implementation in Zambia"

**Supplementary Table 1. Overview of survey, in-depth interview, and focus group participants**

|  | **Survey (n=499)** | | **In-depth Interviews (n=25)** | | **Focus Group Discussions (n=9)** | |
| --- | --- | --- | --- | --- | --- | --- |
|  | **Community Members**  **(n=395)** | **HCWs**  **(n=104)** | **Community Members**  **(n=12)** | **HCWs**  **(n=13)** | **Community Members**  **(n=17)** | **HCWs**  **(n=21)** |
| **Age group** | 30 (23-43) | 33 (28-43) |  |  |  |  |
| 18-24 years | 128 (32.4) | 7 (6.7) | 4 (33.4) | 1 (8.3) | 6 (35.2) | 2 (9.5) |
| 25-29 years | 66 (16.7) | 30 (28.8) | 1 (8.3) | 2 (16.7) | 2 (11.8) | 6 (28.6) |
| 30-34 years | 49 (12.4) | 21 (20.2) | 1 (8.3) | 3 (25) | 1 (5.9) | 6 (28.6) |
| 35-44 years | 66 (16.7) | 26 (25.0) | 1 (8.3) | 4 (33.4) | 5 (29.4) | 4 (19.0) |
| 45-54 years | 37 (9.4) | 14 (13.5) | 3 (25) | 2 (16.7) | 1 (5.9) | 2 (9.5) |
| 55+ years | 49 (12.4) | 6 (5.8) | 2 (16.7) | 0 | 2 (11.8) | 1 (4.8) |
| **Sex** |  |  |  |  |  |  |
| Male | 187 (47.3) | 43 (41.3) | 7 (58.3) | 8 (66.6) | 6 (35.3) | 10 (47.7) |
| Female | 208 (52.7) | 61 (58.7) | 5 (41.7) | 4 (33.4) | 11 (64.7) | 11 (52.3) |
| **Marital Status** |  |  |  |  |  |  |
| Single | 167 (42.3) | 41 (39.4) | 4 (33.3) | - | - | - |
| Stable relationship, not married | 18 (4.6) | 6 (5.8) | 0 | - | - | - |
| Co-habitating/Married | 166 (42.0) | 54 (51.9) | 7 (58.4) | - | - | - |
| Divorced/Separated | 18 (4.6) | 2 (1.9) | 1 (8.3) | - | - | - |
| Widowed | 25 (6.3) | 1 (1.0) | 0 | - | - | - |
| Prefer not to answer | 1 (0.3) | 0 | 0 | - | - | - |
| **Education Level** |  |  |  |  |  |  |
| Less than secondary | 64 (16.2) | 4 (3.8) | 0 | - | - | - |
| Some secondary | 123 (31.1) | 0 | 0 | - | - | - |
| Completed secondary | 156 (39.5) | 27 (26.0) | 10 (83.3) | - | - | - |
| Post-secondary | 52 (13.2) | 73 (70.2) | 2 (16.7) | - | - | - |
| **Employment status** |  |  |  |  |  |  |
| Unemployed | 126 (31.9) | 0 | 3 (25) | 0 | - | 0 |
| Housewife/carer | 34 (8.6) | 0 | 0 | 0 | - | 0 |
| Casual worker/ Piece work | 29 (7.3) | 0 | 4 (33.4) | 0 | - | 0 |
| Self-employed/business | 96 (24.3) | 1 (1.0) | 3 (25) | 0 | - | 0 |
| Formal employee | 42 (10.6) | 65 (62.5) | 1 (8.3) | 7 (53.8) | - | 9 (42.9) |
| Student | 46 (11.6) | 0 | 1 (8.3) | 0 | - | 0 |
| Retired/Volunteer/other | 22 (5.6) | 38 (36.5) | 0 | 6 (46.2) | - | 12 (57.1) |
| **HIV status** |  |  |  |  |  |  |
| Positive | 35 (8.9) | 14 (13.5) | 2 (16.7) | 0 | 0 | 0 |
| Negative | 294 (74.4) | 80 (76.9) | 10 (83.3) | 12 (100) | 16 (94.1) | 0 |
| Unknown | 66 (16.7) | 10 (9.6) | 0 | 0 | 1 (5.9) | 21 (100) |
| **COVID-19 Doses** |  |  |  |  |  |  |
| 0 | 150 (38.0) | 7 (6.7) | 4 | 0 | 4 (23.5) | 0 |
| 1 | 113 (28.6) | 24 (23.1) | 3 | 2 (15.4) | 4 (23.5) | 3 (14.3) |
| 2+ | 132 (33.4) | 73 (70.2) | 5 | 10 (76.9) | 9 (52.9) | 18 (85.7) |
| Unknown | 0 | 0 | 0 | 1 (7.7) | 0 | 0 |

“-“ Indicates that the data was not collected or is not available.

**Supplementary Table 2. Adjusted marginal probability of intention to get a future TB vaccine by subgroup (n=499)**

| **Factor** | **Level** | **Adjusted probability (%)** | **95% CI** | |
| --- | --- | --- | --- | --- |
| **Age** | 18-24 years | 77.2% | 68.7% | 85.6% |
|  | 25-29 years | 83.7% | 75.7% | 91.8% |
|  | 30-34 years | 77.6% | 67.7% | 87.6% |
|  | 35-44 years | 76.8% | 70.0% | 85.6% |
|  | 45-54 years | 74.4% | 62.8% | 86.0% |
|  | 55+ years | 77.4% | 66.5% | 88.3% |
| **Sex at birth** | Male | 75.3% | 69.7% | 80.8% |
|  | Female | 80.6% | 75.9% | 85.4% |
| **Marital status** | Single | 81.8% | 75.5% | 88.0% |
|  | Stable relationship, not married | 77.5% | 61.7% | 93.3% |
|  | Co-cohabiting/Married | 74.2% | 68.3% | 80.1% |
|  | Divorced/Separated | 79.1% | 61.1% | 97.0% |
|  | Widowed | 83.5% | 69.1% | 97.9% |
| **Educational level** | Less than secondary school | 78.5% | 67.8% | 89.3% |
|  | Some secondary school | 82.6% | 75.3% | 90.0% |
|  | Completed secondary school | 76.8% | 70.7% | 82.9% |
|  | Post-secondary school | 75.7% | 67.4% | 84.0% |
| **Employment status** | Unemployed | 77.3% | 69.7% | 85.0% |
|  | Housewife/Carer | 70.8% | 54.7% | 86.9% |
|  | Casual worker/Piece work | 77.8% | 59.2% | 96.4% |
|  | Self-employed/Business | 76.9% | 68.0% | 85.8% |
|  | Formal employee | 82.1% | 73.3% | 91.0% |
|  | Student | 72.2% | 57.5% | 87.0% |
|  | Retired/Volunteer/Other | 83.2% | 73.6% | 93.0% |
| **HCW status** | Community Member | 76.9% | 72.6% | 81.3% |
|  | Healthcare Worker | 82.6% | 72.7% | 92.4% |
| **HIV status** | HIV negative | 76.4% | 72.1% | 80.7% |
|  | HIV positive | 86.2% | 76.8% | 95.7% |
|  | HIV status unknown | 81.4% | 73.0% | 89.7% |
| **TB risk perception*** | Very unlikely | 52.9% | 40.0% | 65.8% |
|  | Somewhat unlikely | 68.1% | 52.9% | 83.4% |
|  | Somewhat likely | 80.8% | 73.3% | 88.2% |
|  | Very likely | 83.3% | 78.8% | 87.7% |
| **Concern about TB severity*** | Not at all concerned | 49.5% | 27.0% | 72.1% |
|  | A little concerned | 57.0% | 41.1% | 73.0% |
|  | Moderately concerned | 75.5% | 65.9% | 85.1% |
|  | Very concerned | 82.4% | 78.5% | 86.3% |
| **Number of COVID-19 doses received** | 0 | 81.8% | 75.6% | 88.1% |
|  | 1 | 72.2% | 65.0% | 79.5% |
|  | 2+ | 79.3% | 73.6% | 85.0% |

*TB risk perception and disease severity concern were not run in the same model due to collinearity. The primary model was run using TB disease severity concern, and the second model was run with TB risk perception.

**Supplementary Table 3. Vaccine information channels and trusted sources among community members and healthcare workers according to stated TB vaccine intention.**

|  | **Overall**  **(n=499)** | **Community Members (n=395)** | | | | **Healthcare workers (n=104)** | | | |
| --- | --- | --- | --- | --- | --- | --- | --- | --- | --- |
|  |  | **Definitely**  **(n=301)** | **Probably**  **(n=79)** | **Probably not**  **(n=10)** | **Definitely not**  **(n=5)** | **Definitely**  **(n=89)** | **Probably**  **(n=13)** | **Probably not**  **(n=1)** | **Definitely not**  **(n=1)** |
| **Current Channels** |  |  |  |  |  |  |  |  |  |
| Television* | 402 (80.6) | 239 (79.4) | 69 (87.3) | 7 (70.0) | 2 (40.0) | 74 (83.2) | 10 (76.9) | 1 (100) | 0 |
| Health Facility | 336 (67.3) | 185 (61.5) | 47 (59.5) | 7 (70.0) | 2 (40.0) | 81 (91.0) | 1 (92.3) | 1 (100) | 1 (100) |
| Social Media | 302 (60.5) | 154 (51.2) | 51 (64.6) | 5 (50.0) | 4 (80.0) | 76 (85.4) | 11 (84.6) | 1 (100) | 0 |
| Radio | 285 (57.1) | 163 (54.2) | 38 (48.1) | 5 (50.0) | 2 (40.0) | 66 (74.2) | 10 (76.9) | 1 (100) | 0 |
| Word of Mouth | 266 (53.3) | 159 (52.8) | 45 (57.0) | 8 (80.0) | 3 (60.0) | 42 (47.2) | 8 (61.5) | 1 (100) | 0 |
| Church | 253 (50.7) | 146 (48.5) | 33 (41.8) | 5 (50.0) | 3 (60.0) | 59 (66.3) | 7 (53.9) | 0 | 0 |
| Workplace | 147 (29.5) | 54 (17.9) | 12 (15.2) | 0 | 1 (20.0) | 69 (77.5) | 10 (76.9) | 1 (100) | 0 |
| Schools | 140 (28.1) | 77 (25.6) | 29 (36.7) | 1 (10.0) | 0 | 28 (31.5) | 5 (38.5) | 0 | 0 |
| Print Media | 86 (17.2) | 37 (12.3) | 7 (8.9) | 1 (10.0) | 1 (20.0) | 33 (37.1) | 6 (46.2) | 1 (100) | 0 |
| **Trusted Sources** |  |  |  |  |  |  |  |  |  |
| Ministry of Health | 437 (87.6) | 259 (86.1) | 68 (86.1) | 10 (100) | 3 (60.0) | 84 (94.4) | 11 (84.6) | 1 (100) | 1 (100) |
| Healthcare workers | 347 (69.5) | 196 (65.1) | 57 (72.2) | 6 (60.0) | 4 (80.0) | 71 (79.8) | 12 (92.3) | 1 (100) | 0 |
| Government | 278 (55.7) | 157 (52.2) | 35 (44.3) | 5 50.0) | 1 (20.0) | 68 (76.4) | 11 (84.6) | 1 (100) | 0 |
| District Health Office | 237 (47.5) | 139 (46.2) | 32 (40.5) | 2 (20.0) | 2 (40.0) | 54 (60.7) | 8 (61.5) | 0 | 0 |
| International Organizations (e.g., WHO) | 141 (28.3) | 69 (22.9) | 19 (24.1) | 2 (20.0) | 2 (40.0) | 42 (47.2) | 7 (53.9) | 0 | 0 |
| Community volunteers | 134 (27.1) | 72 (23.9) | 18 (22.8) | 2 (20.0) | 1 (20.0) | 36 (40.5) | 6 (46.2) | 0 | 0 |
| Vaccine developers | 106 (21.2) | 50 (16.6) | 20 (25.3) | 0 | 1 (20.0) | 29 (32.6) | 6 (46.2) | 0 | 0 |
| Religious leaders | 94 (18.8) | 44 (14.6) | 12 (15.2) | 0 | 2 (40.0) | 31 (34.8) | 5 (38.5) | 0 | 0 |
| Scientists | 93 (18.6) | 44 (14.6) | `12 (15.2) | 1 (10.0) | 1 (20.0) | 27 (30.3) | 8 (61.5) | 0 | 0 |
| Community leaders* | 67 (13.4) | 36 (12.0) | 3 (3.8) | 0 | 2 (40.0) | 25 (28.1) | 1 (7.7) | 0 | 0 |
| Friends and Family | 62 (12.4) | 31 (10.3) | 7 (8.9) | 0 | 2 (40.0) | 17 (19.1) | 5 (38.5) | 0 | 0 |
| Newspapers/magazines | 56 (11.2) | 30 (10.0) | 8 (10.1) | 0 | 1 (20.0) | 15 (16.9) | 2 (15.4) | 0 | 0 |
| Social media* | 45 (9.0) | 17 (5.7) | 9 (11.4) | 1 (10.0) | 2 (40.0) | 12 (13.5) | 4 (30.8) | 0 | 0 |
| Political leaders | 13 (2.6) | 4 (1.3) | 1 (1.3) | 1 (10.0) | 0 | 5 (5.6) | 2 (15.4) | 0 | 0 |
| Traditional healers | 5 (1.0) | 3 (1.0) | 0 | 0 | 0 | 1 (1.1) | 1 (7.7) | 0 | 0 |

*P<0.05 for community members

**Supplementary Table 4. Adjusted marginal probability of reported vaccine information channels and trusted sources**

|  | **High TB vaccine intention**  **(n=390)** | **Lower TB vaccine intention***  **(n=109)** | **Difference in probability for lower TB vaccine intention** |
| --- | --- | --- | --- |
| **Current Channels** |  |  |  |
| Television | 80.6 (76.8-84.4) | 80.4 (72.9-87.9) | -0.2 (-8.6-8.2) |
| Health Facility | 67.9 (63.5-72.2) | 65.4 (56.6-74.2) | -2.5 (-12.5-7.6) |
| Social media | 58.7 (54.5-62.9) | 67.2 (58.9-75.5) | +8.5 (-1.0-18.0) |
| Radio | 58.0 (53.3-62.7) | 53.9 (44.2-63.6) | -4.1 (-15.1-6.9) |
| Word of Mouth | 50.9 (46.1-55.8) | 62.1 (52.3-72.0) | +11.1 (0-22.4) |
| Church | 52.6 (47.8-57.5) | 43.8 (34.6-53.1) | -8.8 (-19.4-1.9) |
| Workplace | 30.0 (26.5-33.5) | 26.8 (19.5-34.2) | -3.2 (-11.4-5.0) |
| Schools | 28.0 (24.2-31.8) | 28.2 (21.3-35.2) | +0.3 (-7.9-8.4) |
| Print Media | 18.0 (14.5-21.5) | 14.5 (8.6-20.5) | -3.5 (-10.5-3.6) |
| **Trusted Sources** |  |  |  |
| Ministry of Health | 88.4 (85.2-91.5) | 84.8 (78.2-91.4) | -3.5 (-10.8-3.8) |
| Healthcare workers | 67.9 (63.5-72.3) | 75.6 (67.4-83.8) | +7.7 (-1.8-17.3) |
| Government | 56.7 (52.1-61.4) | 51.8 (42.6-61.0) | -4.9 (-15.4-5.6) |
| District Health Office | 48.1 (43.3-52.9) | 45.0 (34.8-55.2) | -3.1 (-14.8-8.6) |
| International Organizations (e.g., WHO) | 28.1 (23.9-32.3) | 28.7 (20.5-36.9) | +0.6 (-9.0-10.2) |
| Community volunteers | 26.6 (22.5-30.7) | 29.0 (199.7-38.3) | +2.4 (-7.8-12.7) |
| Vaccine developers | 19.9 (16.1-23.7) | 26.4 (17.8-35.2) | +6.6 (-3.0-16.2) |
| Religious leaders | `18.9 (15.0-2.4) | 19.5 (11.1-27.8) | +0.8 (-8.7-10.3) |
| Scientists | 18.2 (14.5-21.8) | 20.3 (13.0-27.6) | +2.1 (-6.2-10.5) |
| Community leaders | 15.3 (11.8-18.8) | 6.0 (1.3-10.6) | -9.3 (-15.3-3.4) |
| Friends and Family | 11.9 (8.8-14.9) | 14.9 (7.5-22.3) | +3.0 (-5.2-11.2) |
| Newspapers/magazines | 12.0 (8.8-15.2) | 8.8 (3.7-13.9) | -3.2 (-9.4-3.0) |
| Social media | 7.1 (4.9-9.4) | 17.3 (9.3-25.2) | +10.1 (1.6-18.6) |
| Political leaders | 0.2 (0-69.4) | 0.4 (0-1) | +0.1 (-37.0-39.6) |
| Traditional healers | 0.1 (0-22.4) | 14.3 (0-1) | +13.6 (-100, 100) |

*Includes those stating probably will get, probably will not get, definitely will not get

Multivariable model included age, sex, relationship status, education status, role, HIV status, self-reported COVID-19 vaccination doses received, and concern about TB severity

**Supplementary Table 5. Study result implications for TB vaccine-related policy and implementation**

### **Implications for Policy and Implementation**

**Strategies for Early Adopters (External Influence-Driven)**

- **Preparation and Support of HCWs**
- Provide comprehensive, evidence-based training on TB vaccine trial data, safety profiles, and efficacy thresholds so HCWs feel equipped to answer questions.
- Offer practical resources (e.g., transport allowances, adequate staffing, and flexible scheduling) to enable HCWs to serve as effective “vaccine champions” in their communities.

**Strategies for Broader Adoption (Internal Influence-Driven)**

- **Evidence-Based Messaging and Testimonials**
- Emphasize the public health value of partially effective TB vaccines using relatable examples and amplify positive experiences from early adopters, especially HCWs, to boost confidence.

**Strategies Spanning both Adoption Phases**

- **Early Community Engagement**
- Begin sensitization campaigns at least 3–6 months before vaccine availability to preempt rumors and misinformation and build trust and demand.
- Adapt messaging to local languages, cultural contexts, and known social influencers, using a mix of mass media, social media, door-to-door outreach, and community forums.
- **Transparent Policy Communication**
- Stabilize vaccine schedules and communicate clearly about any changes in recommendations or the need for booster doses.
- **Flexible Delivery Strategies**
  - For adults, combine facility-based approaches with community-based strategies at different locations (e.g., churches, markets, and workplaces) to reach diverse populations by minimizing barriers.
  - For adolescents, build on existing school-based initiatives (e.g., HPV vaccination) and incorporate peer-led programs, particularly for out-of-school youth.

re

*These strategies are organized using the Bass Diffusion Model framework,^31^  which distinguishes between early adoption driven by external influences (e.g., formal communication, expert recommendations) and later adoption driven by internal influences (e.g., peer networks, social proof). Strategies in the third section incorporate elements of both, reflecting the interconnected nature of real-world implementation approaches.

**Supplementary Figure 1: Directed acyclic graph for pathogen and intention to receive vaccine indicating a minimal adjustment set including age, community role (healthcare worker or non-healthcare worker community member), household income, and marital status.**

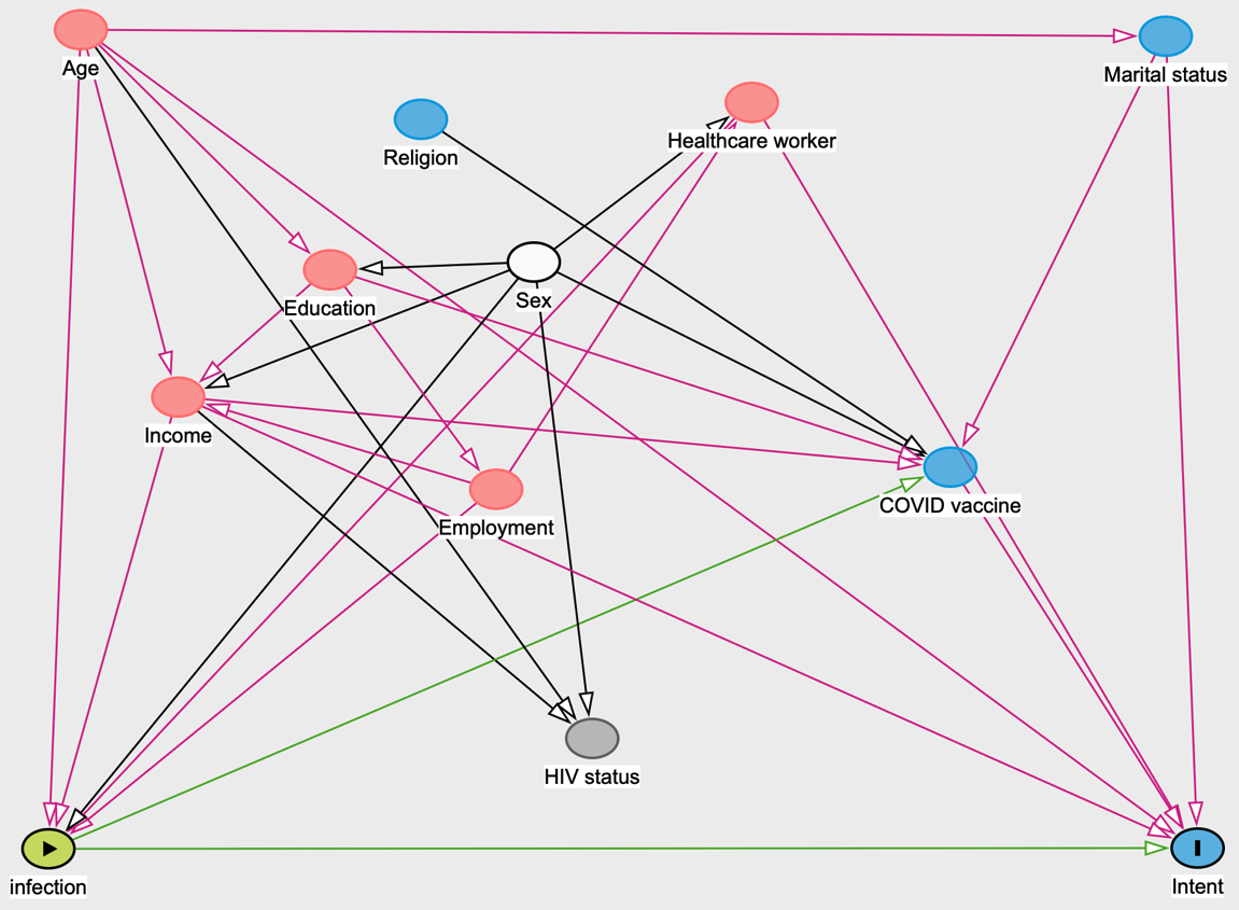

**Supplementary File 1. Standards for Reporting Qualitative Research Checklist**

|  | **Standards for Reporting Qualitative Research (SRQR)*** |  |
| --- | --- | --- |
|  | <http://www.equator-network.org/reporting-guidelines/srqr/> |  |
|  |  | **Page/line no(s).** |
| **Title and abstract** | |  |
|  | **Title** - Concise description of the nature and topic of the study Identifying the study as qualitative or indicating the approach (e.g., ethnography, grounded theory) or data collection methods (e.g., interview, focus group) is recommended | 1 |
|  | **Abstract** - Summary of key elements of the study using the abstract format of the intended publication; typically includes background, purpose, methods, results, and conclusions | 2 |
| **Introduction** | |  |
|  | **Problem formulation** - Description and significance of the problem/phenomenon studied; review of relevant theory and empirical work; problem statement | 3 |
|  | **Purpose or research questio**n - Purpose of the study and specific objectives or questions | 3 |
| **Methods** | |  |
|  | **Qualitative approach and research paradigm** - Qualitative approach (e.g., ethnography, grounded theory, case study, phenomenology, narrative research) and guiding theory if appropriate; identifying the research paradigm (e.g., postpositivist, constructivist/ interpretivist) is also recommended; rationale** | 4-5 |
|  | **Researcher characteristics and reflexivity** - Researchers’ characteristics that may influence the research, including personal attributes, qualifications/experience, relationship with participants, assumptions, and/or presuppositions; potential or actual interaction between researchers’ characteristics and the research questions, approach, methods, results, and/or transferability | 6 |
|  | **Context** - Setting/site and salient contextual factors; rationale** | 4 |
|  | **Sampling strategy** - How and why research participants, documents, or events were selected; criteria for deciding when no further sampling was necessary (e.g., sampling saturation); rationale** | 4 |
|  | **Ethical issues pertaining to human subjects** - Documentation of approval by an appropriate ethics review board and participant consent, or explanation for lack thereof; other confidentiality and data security issues | 4-5 |
|  | **Data collection methods** - Types of data collected; details of data collection procedures including (as appropriate) start and stop dates of data collection and analysis, iterative process, triangulation of sources/methods, and modification of procedures in response to evolving study findings; rationale** | 5 |
|  | **Data collection instruments and technologies** - Description of instruments (e.g., interview guides, questionnaires) and devices (e.g., audio recorders) used for data collection; if/how the instrument(s) changed over the course of the study | 5 |
|  | **Units of study** - Number and relevant characteristics of participants, documents, or events included in the study; level of participation (could be reported in results) | 6 |
|  | **Data processing** - Methods for processing data prior to and during analysis, including transcription, data entry, data management and security, verification of data integrity, data coding, and anonymization/de-identification of excerpts | 5-6 |
|  | **Data analysis** - Process by which inferences, themes, etc., were identified and developed, including the researchers involved in data analysis; usually references a specific paradigm or approach; rationale** | 5-6 |
|  | **Techniques to enhance trustworthiness** - Techniques to enhance trustworthiness and credibility of data analysis (e.g., member checking, audit trail, triangulation); rationale** | 4-6 |
| **Results/findings** | |  |
|  | **Synthesis and interpretation** - Main findings (e.g., interpretations, inferences, and themes); might include development of a theory or model, or integration with prior research or theory | 6-21 |
|  | **Links to empirical data** - Evidence (e.g., quotes, field notes, text excerpts, photographs) to substantiate analytic findings | 6-21 |
| **Discussion** | |  |
|  | **Integration with prior work, implications, transferability, and contribution(s) to the field -** Short summary of main findings; explanation of how findings and conclusions connect to, support, elaborate on, or challenge conclusions of earlier scholarship; discussion of scope of application/generalizability; identification of unique contribution(s) to scholarship in a discipline or field | 22-24 |
|  | **Limitations** - Trustworthiness and limitations of findings | 23-24 |
| **Other** | |  |
|  | **Conflicts of interest** - Potential sources of influence or perceived influence on study conduct and conclusions; how these were managed | 25 |
|  | **Funding** - Sources of funding and other support; role of funders in data collection, interpretation, and reporting | 25 |
|  | *The authors created the SRQR by searching the literature to identify guidelines, reporting standards, and critical appraisal criteria for qualitative research; reviewing the reference lists of retrieved sources; and contacting experts to gain feedback. The SRQR aims to improve the transparency of all aspects of qualitative research by providing clear standards for reporting qualitative research. |  |
|  | **The rationale should briefly discuss the justification for choosing that theory, approach, method, or technique rather than other options available, the assumptions and limitations implicit in those choices, and how those choices influence study conclusions and transferability. As appropriate, the rationale for several items might be discussed together. |  |
|  | **Reference:** |  |
|  | O'Brien BC, Harris IB, Beckman TJ, Reed DA, Cook DA. **Standards for reporting qualitative research: a synthesis of recommendations.** *Academic Medicine*, Vol. 89, No. 9 / Sept 2014  DOI: 10.1097/ACM.0000000000000388 |  |

**Supplementary File 2. Disease Information Sheet Reviewed with Participants Before Survey Completion**

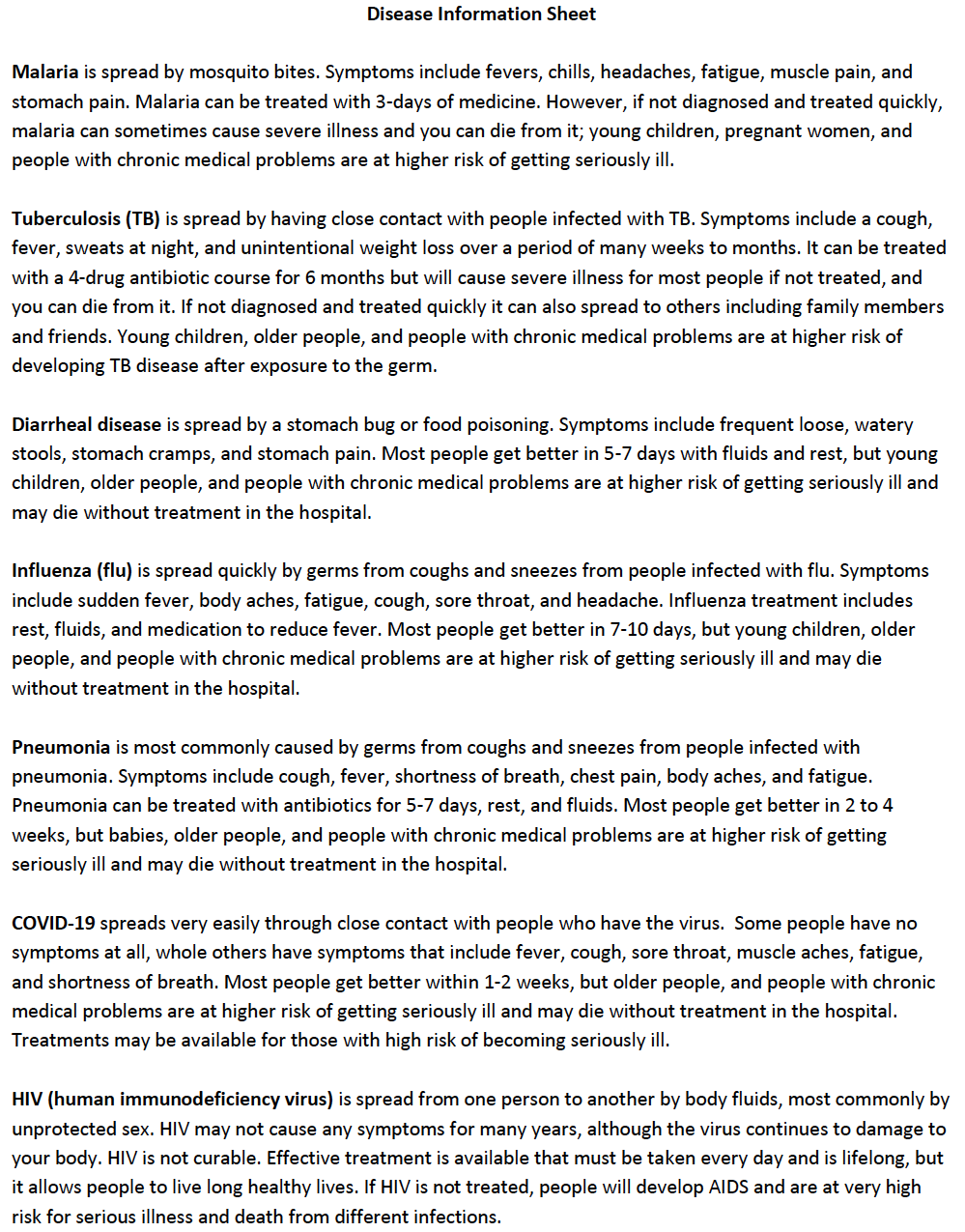
